# Surgical and percutaneous coronary revascularization in patients with multivessel or left main disease; what happens beyond five years? A systematic review and study level meta-analysis of randomized trials

**DOI:** 10.1101/2023.03.17.23287425

**Authors:** Francesco Formica, Daniel Hernandez-Vaquero, Domenico Tuttolomondo, Alan Gallingani, Gurmeet Singh, Claudia Pattuzzi, Giampaolo Niccoli, Roberto Lorusso, Francesco Nicolini

**Affiliations:** University of Parma, Department of Medicine and Surgery, Parma, Italy; Cardiac Surgery Department, Hospital Universitario Central de Asturias, Oviedo, Spain; Cardiology Unit, University Hospital of Parma, Parma, Italy; Cardiac Surgery Unit, University Hospital of Parma, Parma, Italy; Department of Critical Care Medicine and Division of Cardiac Surgery, Mazankowski Alberta Heart Institute, University of Alberta, Edmonton, Canada; Cardio-Thoracic Department, Maastricht University Medical Centre, Heart and Vascular Centre, Maastricht, The Netherlands; Cardiovascular Research Institute Maastricht (CARIM), Maastricht, The Netherlands

**Keywords:** coronary artery bypass grafting, percutaneous coronary intervention, drug-eluting stent, survival, meta-analysis, Long-term follow-up, extended follow-up, randomized trials

## Abstract

Meta-analysis exploring outcomes beyond 5-years of trials comparing coronary artery bypass graft (CABG) with percutaneous coronary intervention (PCI) utilizing drug-eluting stents in patients with coronary artery disease (CAD), are missing. We conducted a meta-analysis to compare very long-term outcomes, between the two interventions.

Using electronic databases, we retrieved 4 trials, between January, 2010 and January, 2023. The primary endpoint was all-cause mortality. Kaplan-Meier curves of endpoint was reconstructed. Comparisons were made by Cox-linear regression frailty model and by landmark analysis. A flexible parametric model for survival analysis was used to obtain the time-dependent hazard-ratio. A random-effect method was applied.

5180 patients were included and randomized to CABG (n=2586) or PCI with DES (n=2594). During 10-year follow-up, PCI showed an overall higher incidence of all-cause mortality [hazard ratio (HR) 1.19; 95% confidence interval (CI), 1.104-1.32; p=0.008)]. At landmark analysis, PCI showed higher risk of the endpoint within 5-years (HR 1.2; 95% CI, 1.06-1.53; p=0.008) while no difference was found at 5–10-year period (HR,1.03; 95%CI, 0.84-1.26; p=0.76). The time-varying HR analysis of PCI versus CABG was consistent with the results of the landmark analysis. There was no long-term difference between the two interventions for myocardial infarction (MI) (OR,1.42; 95%CI, 0.92-2.18; p=0.11), composite of all-cause mortality, stroke or MI (OR,1.07; 95%CI, 0.84-1.36; p=0.57), stroke (OR,0.97; 95%CI, 0.59-1.59; p=0.91) and cardiovascular death (OR,1.02; 95%CI, 0.75-1.40; p=0.90), while PCI was associated with an increased risk for repeat revascularization (OR,2.11; 95%CI, 1.58-2.81; p<0.001) and major adverse cardiac and cerebrovascular events (OR,1.41; 95%CI, 1.13-1.75; p<0.0001). In conclusion, in patients with CAD, there was a significantly overall higher incidence of all-cause mortality after PCI compared to CABG beyond 5-year follow-up. Specifically, CABG is still favorable beyond 5 years and maintains its gold standard role for the CAD treatment; PCI has an evident higher mortality during the first 5 years and a comparable outcome beyond 5 years.

## Introduction

The 2018 European and 2021 American Guidelines recommended percutaneous coronary intervention (PCI) as an appropriate coronary revascularization strategy as alternative to coronary artery bypass grafting (CABG) in patients with left main disease (LMD) and multivessel disease (MVD), as well as with low-intermediate coronary complexity (1,2). These recommendations are based on 5-year follow-up results of randomized clinical trials (RCTs) comparing PCI with drug-eluting stents (DES) and CABG, published during the last 10 years. However, life expectancy for a 80-year person of the general population in Europe and USA is around 9 years (3,4). Therefore, 5 years results are clearly not enough to decide the best strategy for the vast majority of patients. Recent meta-analyses of RCTs comparing PCI with DES and CABG in patient with LMD and or MVD, have reported conflicting results between the two interventions in terms of 5-year overall survival, stroke, myocardial infarction (MI) and repeat revascularization, although most of pooled results showed an advantage in favour of CABG over PCI (5–7). However, the choice of the optimal mode of revascularization remains controversial especially for many patients that have a life expectancy of more than 10 years.

End-points of many RCTs are limited at 5 years of follow-up. But survival curves of these RCTs frequently provide longer information. This information is not usually useful in individual studies due to low statistical power after 5 years because of deaths and censored events. However, this low statistical power could be overcome by pooled analysis, which is one of the aims of the meta-analysis (8).

Therefore, given the importance of the optimal revascularization strategy is still under debate, and considering that there are no meta-analyses of RCTs exploring the results of PCI with DES and CABG beyond 5 years, we conducted a comprehensive systematic review and meta-analysis with the aim of comparing very long-term outcomes, beyond 5-years, between the two interventions.

## Methods

As the aggregated data were extracted from the published articles included in the analysis, this meta-analysis is exempted from Ethical Committee (EC) evaluation as the investigators of each trial obtained the approval from the local Ethical Committees (EC). The meta-analysis adhered to the Preferred Reporting Items for Systematic Reviews and Meta-Analyses (PRISMA) guidelines (9) and the study protocol was registered and published online in PROSPERO (The International Prospective Register of Systematic Reviews; ID: CRD42023401293).

### Search strategy

The search strategy consisted of a comprehensive review of relevant studies published between January 1, 2010 and January 31, 2023 in three electronic databases, PubMed, Cochrane Central Register of Controlled Trials (CENTRAL) and EMBASE. The references list of previous meta-analyses and relevant articles were also used to complete the search.

Using Boolean operators (“AND” or “OR”), the search strings included (‘multivessel coronary artery disease’) AND (‘left main disease’ OR ‘left main coronary artery disease’) AND (‘coronary artery bypass’ OR ‘CABG’) AND (‘percutaneous coronary intervention’ OR ‘PCI’) AND (‘drug-eluting stents’ OR ‘DES’ OR stenting) AND (‘randomized’ OR ‘randomised’ OR ‘trials’) AND (‘long-term follow-up’ OR ‘extended follow-up’). A medical librarian refined the literature search. The search algorithm is reported in Supplementary Material (Table S1).

### Inclusion criteria

Study eligibility criteria followed the PICOS format (Population; Intervention; Comparison; Outcomes; Studies). Population: patients with CAD affected by LMD and or MVD and deemed eligible for either CABG or PCI; Intervention: PCI; Comparison: CABG operation; Outcomes: overall survival and incidence of stroke, MI and repeat revascularization at longest follow-up; Studies: only RCTs written in English languages that reported graphed Kaplan–Meier (KM) curves of very long-term survival (beyond 5-year follow-up) of the outcomes of interest. Two authors (DT, CP) independently scanned and reviewed titles and abstracts and disagreement was resolved by a senior author (FF).

### Data extraction and collection

Two authors (AG, DT) independently extracted data from main text and supplementary materials of the RCTs included in the analysis. Data were then collected in a standard table sheet database (Microsoft Office Excel 2016, Microsoft, Redmond, WA, USA). The included trials were listed by first author, study period and year of publication, preoperative characteristics and postoperative outcomes. Disagreement was solved by a senior author (FF).

### Risk of bias assessment

Two authors (NF, FF) assessed the quality of the studies and the risk of bias using the Cochrane Collaboration revised tool for randomized control trials (RoB 2) (9) (Supplementary Table S2).

### Endpoints

The primary endpoint was the incidence of all-cause mortality. The secondary endpoints were repeat coronary revascularization, myocardial infarction (MI), incidence of cardiovascular (CV) death, stroke, composite outcomes (all-cause mortality, stroke and MI) and the incidence of major adverse cardiac and cerebrovascular events (MACCE) including all-cause mortality, stroke, MI and repeat coronary revascularization.

### Statistical analysis

#### Data pooling and meta-analysis

Continuous variables were reported as mean ± standard deviation (SD). Variables expressed in median and interquartile ranges were converted into mean and SD using a validated formula (11). Categorical variables were reported as number and percentages.

Primary endpoint was analyzed with two-stage approach to conduct the individual patient data (IPD) reconstruction meta-analysis (12). Secondary outcomes were compared using odd ratio (OR) and 95% confidence interval (CI) and effect estimates were calculated according to the random-effect model and the DerSimonian-Laird method and were represented with the forest plot. An OR > 1 indicated an advantage of CABG arm over PCI. Heterogeneity was evaluated with chi-squared and I^2^ tests and defined as absent or low for I^2^ ranging from 0% to 25%, moderate for I^2^ ranging from 26% to 50% and high for I^2^ above 50% (13).

#### Individual patient data reconstruction

To assess the entire follow-up duration of each trial we performed an IPD analysis applying an iterative approach to reconstruct the original database of each trial using the method described by Wei, Royston et al. (14,15). We employed a dedicated software (GetData Graph Digitizer version 2.5.3; http://getdata-graph-digitizer.com) to digitize the K-M curves by importing the time (abscissa-x) and survival probability (ordinate-y) values from the original K-M curves. The databases were reconstructed by combining the extracted value of time and survival with the patients at risk for the time points indicated in the original K-M curve. Quality assessment of the databases were performed by visually comparing the K-M curves and the estimated patients at risk from the reconstructed databases with the original ones. Then, we merged data of reconstructed databases in a single database to recalculate the aggregated survival curves and the life tables. By doing this, we simulated a patient-level meta-analysis (16).

#### First stage of two-stage approach meta-analysis

Cox proportional hazards models with inclusion of frailty term to account for heterogeneity amongst trials was used to compare the two arms and the hazard ratio (HR) with 95% CIs were calculated and reported. The proportional of hazard assumption (PHA) was assessed for the primary endpoint and was tested by visual inspection of K-M curves, log-minus-log plots, predicted versus observed survival curves and the scaled Schoenfeld residuals. A p<0.05 indicated a violation of proportionality.

Given the potential different long-term (0-5 years) and very long-term risks (5-10 years) of the two revascularization approaches, landmark analysis with a 5-years cut-off was planned in the statistical analysis and performed.

Further, a flexible parametric model for survival analysis was used to obtain the time-dependent HRs (Royston-Parmar models) using a restricted cubic spline function. The baseline cumulative hazard was modeled using spline with 5 degrees of freedom (df; 4 intermediate knots and two knots at each boundary); the time-varying effect of intervention was modelled using a restricted cubic spline with 3 df (2 internal knots).

#### Second stage of two-stage meta-analysis

In the second stage we assessed the treatment effect estimate at longest available follow-up as OR with its 95% CI. An OR > 1 indicated an advantage of CABG arm. Effect measure was calculated according to the random-effect model and the DerSimonian-Laird method and was represented with the forest plot. Heterogeneity was evaluated with chi-squared and I^2^ tests and defined as early described (13).

#### Sensitivity analysis

As a sensitivity analysis, we compared the pooled HRs and 95% CIs of both one-stage and two-stage analysis and then, to increase the accuracy of our results, we performed an extra sensitivity analysis according to the leave-one-out method (17) to identify the influence of a single study on the primary outcome, if heterogeneity was significant.

#### Restricted mean survival time

We used the restricted mean survival time (RMST) method to compare the mean survival time between CABG and PCI at a specified truncation time (*t\**). The RMST represents a measure of life expectancy between the time of intervention and the *t\** and was calculated as the area under the survival curve for each arm. We selected *t\** = 5-years, *t\** = 8-years and *t\** = 10-years for the following reasons: first, all trials have follow-up longer than 5 years; second, recent meta-analyses reported survival results at 5-years follow-up; third, the 8-years was the longest follow-up shared by the four trials; last, 10 years was the longest follow-up available. We calculated the difference of RMST between CABG and PCI (rmstD = rmstCABG - rmstPCI), which is interpretable as the number of life-years gained with PCI compared to CABG (18,19). All the statistical analyses were computed with Stata/SE version 16.1 (Stata Corp, College Station, Tex). A two-tailed p-value <0.05 indicated statistical significance.

## Results

The literature search identified 475 records and 13 studies were considered relevant and then retrieved. Among them, 4 RCTs [BEST (NCT05125367 and NCT00997828) FREEDOM (NCT00086450), PRECOMBAT (NCT03871127 and NCT00422968) and SYNTAX (NCT03417050) (20–23)] met the eligibility criteria and were included in the final analysis. The PRISMA Flow Chart of study selection is shown in Supplemental Figure 1. The trials included 5180 patients, including 2586 patients randomized to CABG and 2594 patients to PCI with DES. All trials presented follow-up beyond 5-years with a mean weighted follow-up duration of 10.23 years. Specifically, BEST, PRECOMBAT and SYNTAX trials have reported 10-year follow-up for all-cause mortality, while FREEDOM trial has reported 8-year follow-up. In addition, FREEDOM trial reported separately survival analysis of whole cohort (n=1900) and of extended cohort (n=943). Baseline variables of patients enrolled in each trial are reported in Table 1. Baseline characteristics of single trials are showed in Table 2. The endpoint definition of single trials is presented in supplementary material (Table S3).

**Table 1.** Baseline variables of enrolled patients.

|  | BEST |  |  |  | FREEDOM<sup>a</sup> |  |  |  | PRECOMBAT |  |  |  | SYNTAX |  |  |  |
| --- | --- | --- | --- | --- | --- | --- | --- | --- | --- | --- | --- | --- | --- | --- | --- | --- |
|  | PCI |  | CABG |  | PCI |  | CABG |  | PCI |  | CABG |  | PCI |  | CABG |  |
| No. of patients | 438 |  | 442 |  | 475/953 |  | 482/947 |  | 300 |  | 300 |  | 903 |  | 897 |  |
|  | Mean or no. | SD or % | Mean or no. | SD or % | Mean or no. | SD or % | Mean or no. | SD or % | Mean or no. | SD or % | Mean or no. | SD or % | Mean or no. | SD or % | Mean or no. | SD or % |
| Age | 64 | 9,3 | 64,9 | 9,4 | 62,9 | 9,3 | 63,1 | 9,4 | 61,8 | 10 | 62,7 | 9,5 | 65,2 | 9,7 | 65 | 9,8 |
| Male sex | 304 | 69,4 | 325 | 73,5 | 361 | 76 | 344 | 71,4 | 228 | 76 | 231 | 77 | 690 | 76,4 | 708 | 78,9 |
| BMI | 24,7 | 2,9 | 25 | 2,9 | 29,7 | 5,2 | 29,9 | 5,4 | 24,6 | 2,7 | 24,5 | 3 | 28,1 | 4,8 | 27,9 | 4,5 |
| Medical diabetes |  |  |  |  |  |  |  |  |  |  |  |  |  |  |  |  |
| *Any* | 177 | 40,4 | 186 | 42,1 | 953 | 100 | 947 | 100 | 102 | 34 | 90 | 30 | 231 | 25,6 | 221 | 24,6 |
| *Requiring Insuline* | 20 | 84,6 | 18 | 4,1 | 322 | 33,8 | 293 | 947 | 10 | 3,3 | 9 | 3 | 89 | 9,9 | 93 | 10,4 |
| Hypertension | 296 | 67,6 | 295 | 66,7 | 411 | 86,5 | 407 | 84,4 | 163 | 54,3 | 154 | 51,3 | 630 | 69,8 | 574 | 64 |
| Hyperlipidemia | 239 | 54,6 | 222 | 50,2 | -- | -- | -- | -- | 127 | 4,3 | 120 | 40 | 711 | 78,7 | 692 | 77,2 |
| Smoker (current) | 88 | 20,1 | 89 | 20,1 | 80 | 16,8 | 82 | 17 | 89 | 29,7 | 83 | 27,7 | 167 | 18,5 | 197 | 22 |
| Previous PCI | 30 | 6,8 | 38 | 8,6 | -- | -- | -- | -- | 38 | 12,7 | 38 | 12,7 | -- | -- | -- | -- |
| Previous MI | 25 | 5,7 | 29 | 6,6 | 109 | 22,9 | 96 | 19,9 | 13 | 4,3 | 20 | 6,7 | 288 | 31,9 | 303 | 33,8 |
| Previous CHF | 16 | 3,7 | 12 | 2,7 | -- | -- | -- | -- | 0 | 0 | 2 | 0,7 | 36 | 4 | 47 | 5,3 |
| Previous stroke | 37 | 8,4 | 33 | 7,5 | 25 | 5,7 | 21 | 4,4 |  |  |  |  | 35 | 3,9 | 43 | 4,8 |
| Chronic Renal failure | 9 | 2,1 | 7 | 1,6 | -- | -- | -- | -- | 4 | 1,3 | 1 | 0,3 |  |  |  |  |
| PVD | 15 | 3,4 | 12 | 2,7 | -- | -- | -- | -- | 15 | 5 | 7 | 2,3 | 73 | 8,1 | 75 | 8,4 |
| COPD | 8 | 1,8 | 6 | 1,4 | -- | -- | -- | -- | 6 | 2 | 10 | 3,3 |  |  |  |  |
| Clinical presentation |  |  |  |  |  |  |  |  |  |  |  |  |  |  |  |  |
| *Stable angina* | 210 | 47,9 | 204 | 46,2 | -- | -- | -- | -- | 160 | 53,3 | 137 | 45,7 | 514 | 56,9 | 513 | 57,2 |
| *Unstable angina* | 185 | 42,2 | 199 | 45 | -- | -- | -- | -- | 128 | 42,7 | 144 | 48 | 261 | 28,9 | 251 | 28 |
| *AMI (<90 days)* | 43 | 9,8 | 39 | 8,8 | -- | -- | -- | -- | -- | -- | -- | -- | -- | -- | -- | -- |
| *Recent MI (within 7 days of random)* | -- | -- | -- | -- | -- | -- | -- | -- | -- | -- | -- | -- | -- | -- | -- | -- |
| *Recent acute coronary syndrome* | -- | -- | -- | -- | 161 | 33,9 | 152 | 31,5 | -- | -- | -- | -- | -- | -- | -- | -- |
| *Unstable angina and recent NSTEMI* | -- | -- | -- | -- | -- | -- | -- | -- | 12 | 4 | 19 | 6,3 | -- | -- | -- | -- |
| EF | 59,1 | 8,5 | 59,9 | 8,1 | 65,7 | 12,1 | 66,6 | 10,5 | 61,7 | 8,3 | 60,6 | 8,5 | -- | -- | -- | -- |
| N of diseased vessels |  |  |  |  |  |  |  |  |  |  |  |  |  |  |  |  |
| 3 | 330 | 75,3 | 349 | 79 | 394/474 | 83,1 | 414/480 | 86,3 | 122 | 40,7 | 123 | 41 | 546 | 60 | 549 | 61 |
| 2 | 108 | 24,7 | 93 | 21 | -- | -- | -- | -- | 101 | 33,7 | 90 | 30 | -- | -- | -- | -- |
| EuroSCORE |  |  |  |  |  |  |  |  |  |  |  |  |  |  |  |  |
| mean score | 2,9 | 2 | 3 | 2,1 | 2,8 | 2,7 | 2,8 | 2,8 | 2,6 | 1,8 | 2,8 | 1,9 | 2,8 | 2,6 | 3,8 | 2,7 |
| median |  |  |  |  | 2,0 | (1,3-3,2) | 2,1 | (1,3-3,3) | -- | -- | -- | -- | -- | -- | -- | -- |
| SYNTAX score |  |  |  |  |  |  |  |  |  |  |  |  |  |  |  |  |
| mean score | 24,2 | 7,5 | 24,6 | 8,1 | 26,9 | 8,2 | 26,1 | 8,1 | 24,3 | 9,6 | 25,3 | 10,9 | 28,4 | 11,5 | 29,1 | 11,4 |
| median | -- | -- | -- | -- | 27 | (21-31,5) | 26 | (20-31) | -- | -- | -- | -- | -- | -- | -- | -- |
| >=33 | 66 | 15,1 | 79 | 17,9 | 98 | 20,6 | 92/479 | 19,2 | 58 | 19,3 | 68 | 22,7 | 290 | 32,1 | 315 | 35,1 |
| 22-32 | 187 | 42,7 | 177 | 40,0 | 228 | 48 | 220/479 | 45,9 | 102 | 34,0 | 97 | 32,3 | 310 | 34,3 | 300 | 33,4 |
| <=22 | 185 | 42,2 | 186 | 42,5 | 149 | 31,4 | 167/479 | 34,9 | 129 | 43,0 | 104 | 34,7 | 299 | 33,1 | 275 | 30,7 |

a) FREEDOM trial data refer to whole cohort; PCI, percutaneous coronary intervention; CABG, coronary artery bypass grafting; BMI, body mass index; MI, myocardial infarction; CHF, congestive heart failure; PVD, peripheral vascular disease; COPD; chronic obstructive pulmonary disease; EF, ejection fraction.

**Table 2.**
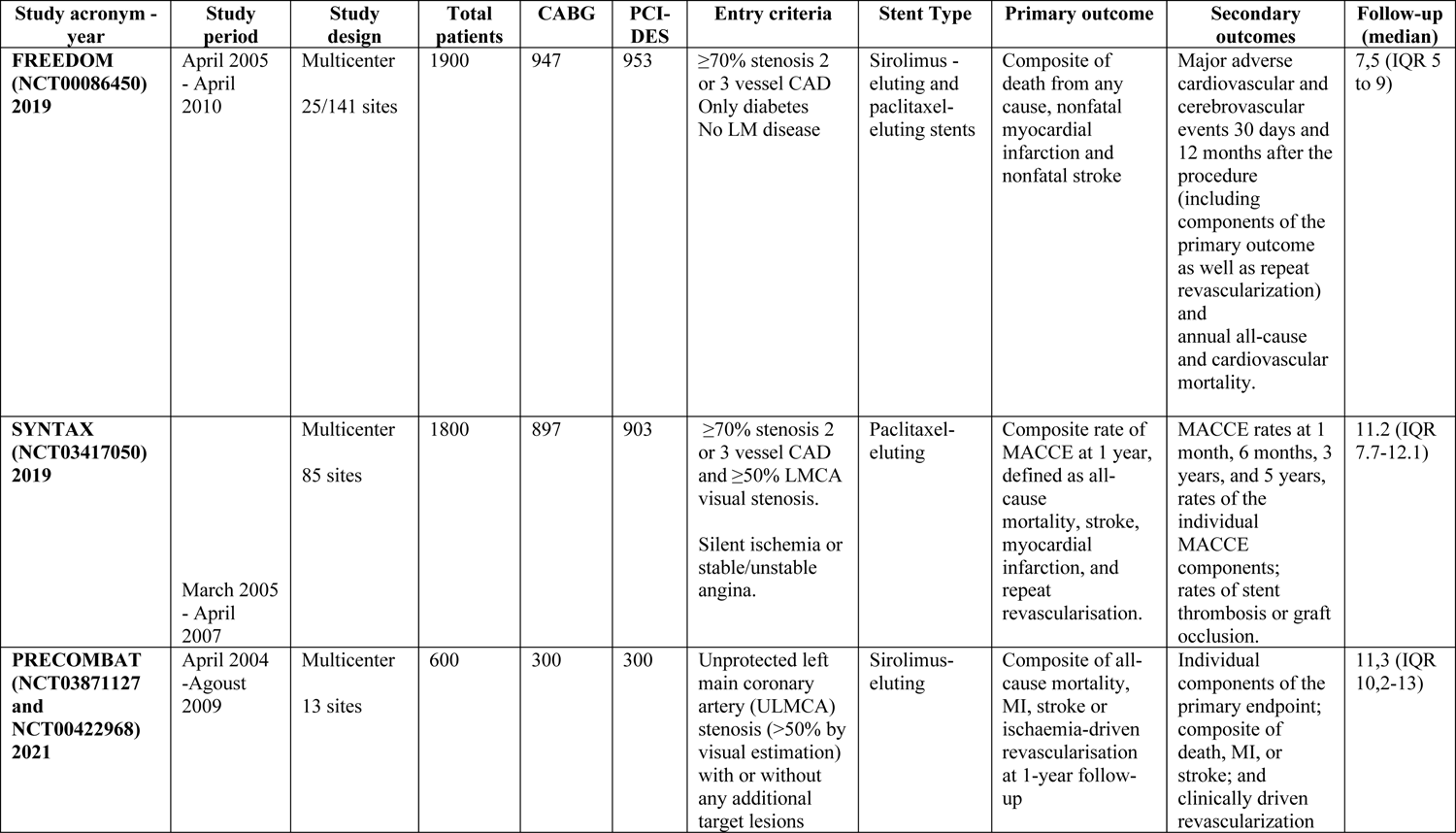
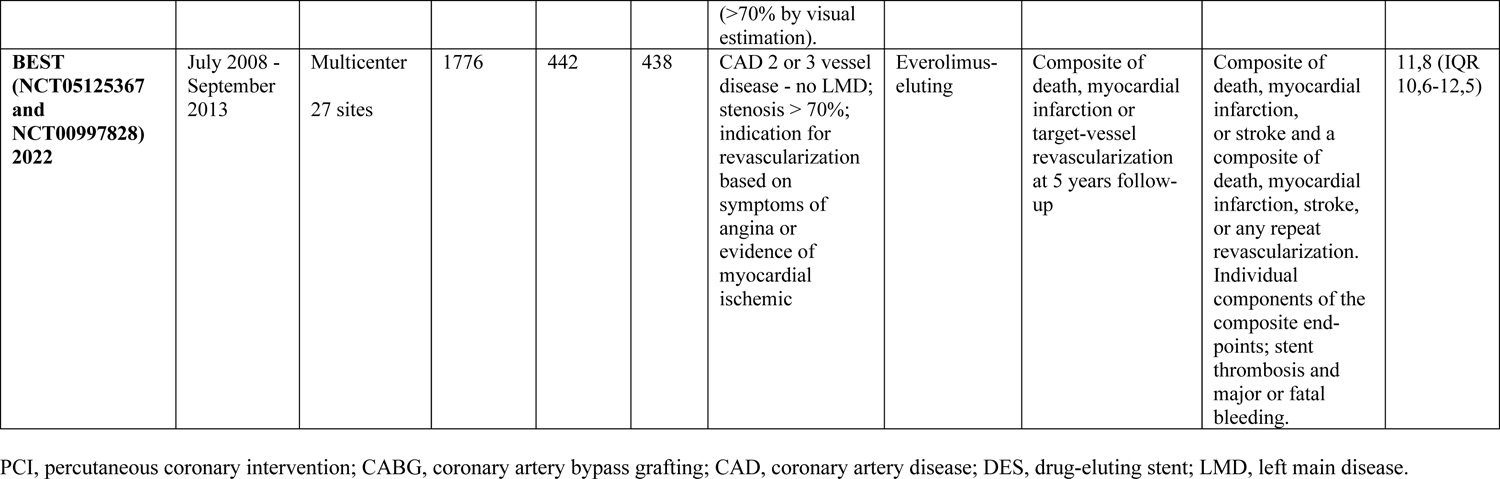
Baseline characteristics of trials included in the meta-analysis

## Primary endpoints

### All-cause mortality

The Cox linear regression frailty model revealed that PCI was associated with a higher rate of all-cause mortality compared to CABG (HR 1.19; 95% CI, 1.04-1.32; p = 0.008, frailty theta 0.08) (Fig. 1A). The PHA was not violated (p=0.4). Additional log-minus-log survival curves, predicted-versus-observed survival functions and the scaled Schoenfeld residuals plot are provided in the supplemental materials (Fig S2 to S4). The landmark analysis showed a greater risk of adverse event for PCI compared to CABG at 0–5-year period (HR 1.2; 95% CI, 1.06-1.53; p = 0.008), while no difference was found at 5–10-year period (HR 1.03; 95% CI, 0.84-1.26; p = 0.76) (Fig. 1B). We observed a statistically significant 5-year RMST difference of 0.07 years (25 days) (95% CI, 0.01-0.125; p=0.02) gained with CABG. After 8 years of follow-up, the RMST difference was statistically significant at 0.14 years (1.7 months) (95% CI, 0.03-0.25; p=0.009) gained with CABG. The 10-year RMST difference was statistically significant at 0.20 years (95% CI, 0.05-0.35; p=0.007), suggesting a prolonged life expectancy by 0.20 years (2.4 months) in patients with CABG, compared to patients with PCI (Fig. 1A and Supplementary Table S4).

**Figure 1.**
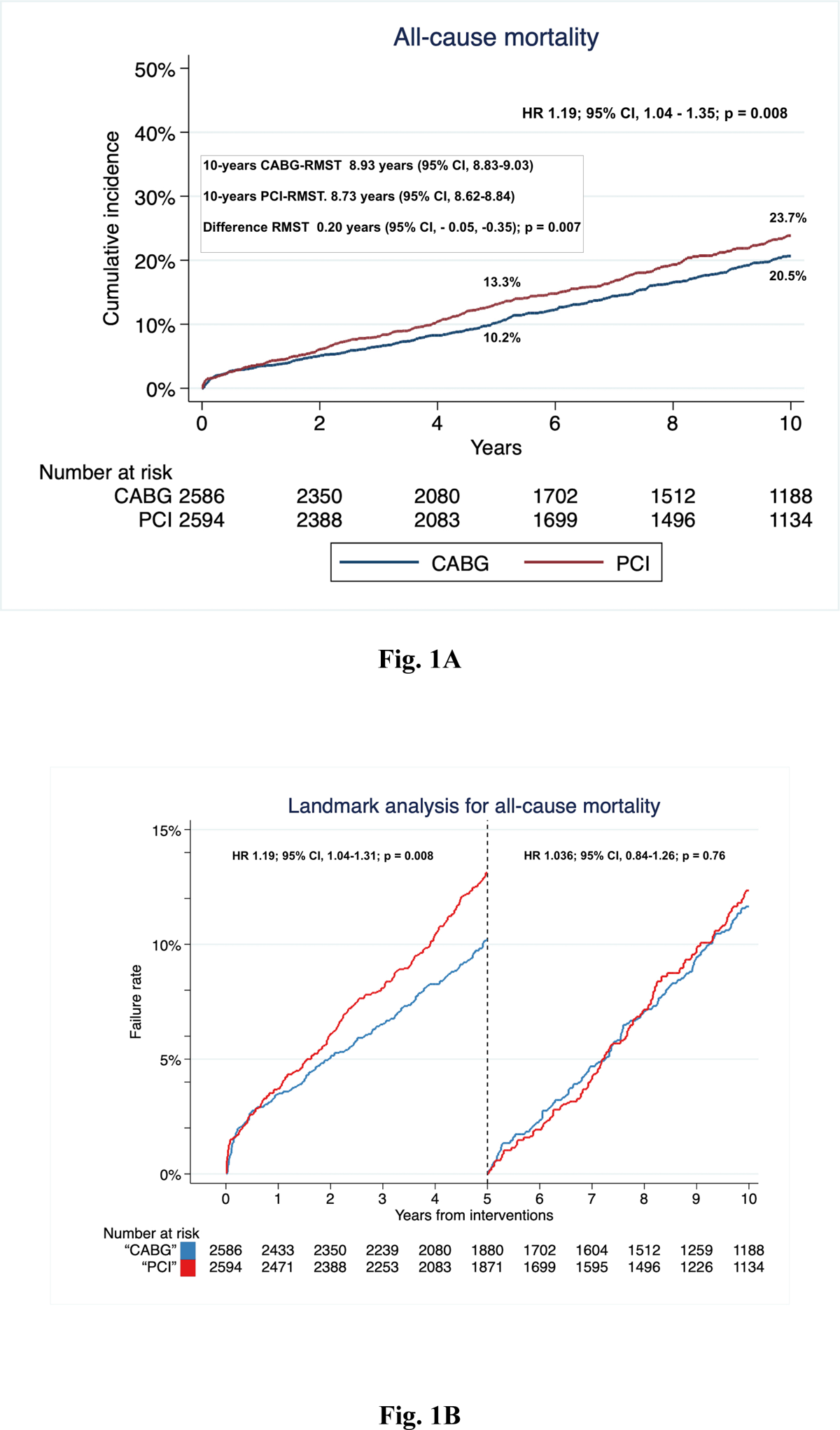
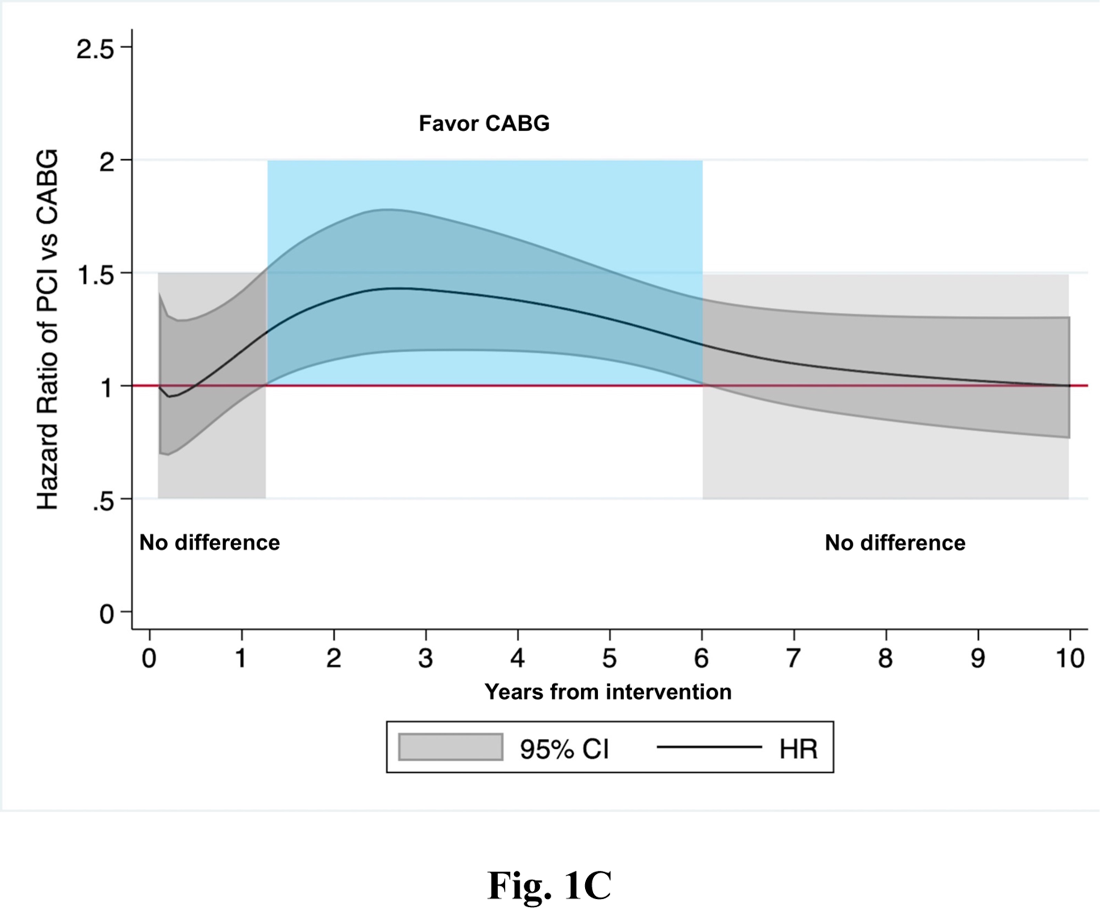
**(B)** Kaplan–Meier incidence function plot of reconstructed individual patient data analysis for all-cause mortality following coronary artery bypass grafting (CABG) or percutaneous coronary intervention (PCI) and 10-year restricted mean survival time (RMST). **(B)** Landmark analysis for all-cause mortality following CABG or PCI. **(C)** Hazard ratio trend over time for all-cause CABG versus PCI estimated by fully parametric model for survival analysis. *HR, hazard ratio*; *OR, odd ratio; CI, confidence interval*.

The time-varying HR analysis of PCI versus CABG was consistent with the results of the landmark analysis (Fig. 1C). PCI and CABG showed comparable results in the first year after surgery. Thereafter, the benefit of CABG became clearly superior to PCI until about 6 years. Beyond 6-years the benefit of CABG was lost and the two interventions were comparable. In the cumulative two-stage meta-analysis, the point estimate for all-cause mortality at the longest follow-up showed a higher risk of death for PCI compared with CABG (OR 1.24, 95% CI, 1.06–1.45; p = 0.01) with low heterogeneity (I^2^=14.37%) (Fig. 2).

**Figure 2.**
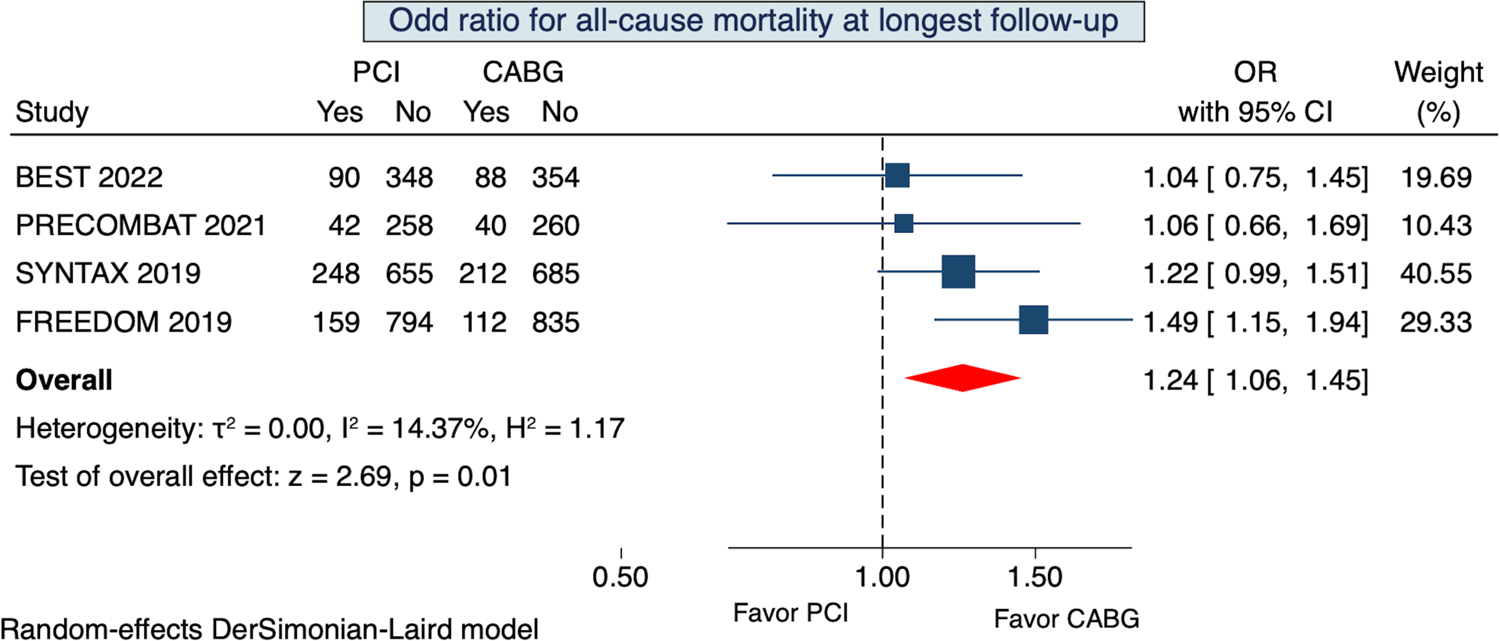
**(A)** Overall effect for all-cause mortality at longest follow-up following coronary artery bypass grafting (CABG) or percutaneous coronary intervention (PCI). *OR, odd ratio; CI, confidence interval;*

We performed both first-stage and second-stage analysis of two-stage approach analysis including data of extended follow-up FREEDOM trial. Results of the Cox regression linear frailty model (HR 1.16; 95% CI, 1.01-1.32; p = 0.03, frailty theta 0.08) and the cumulative meta-analysis (OR 1.23; 95% CI, 1.06-1.42; p = 0.01) were consistent with the main analisys.

### Secondary endpoints

#### Myocardial infarction

Data on MI beyond 5-year follow-up were reported in BEST, FREEDOM and PRECOMBAT trials. In the FREEDOM trial, data on MI were recorded in 415 patients. The overall effect for MI showed that PCI and CABG were comparable (OR 1.42; 95% CI, 0.92-2.18; p=0.11) with no statistical heterogeneity (I^2^=0%) (Fig. 3A).

**Figure 3.**
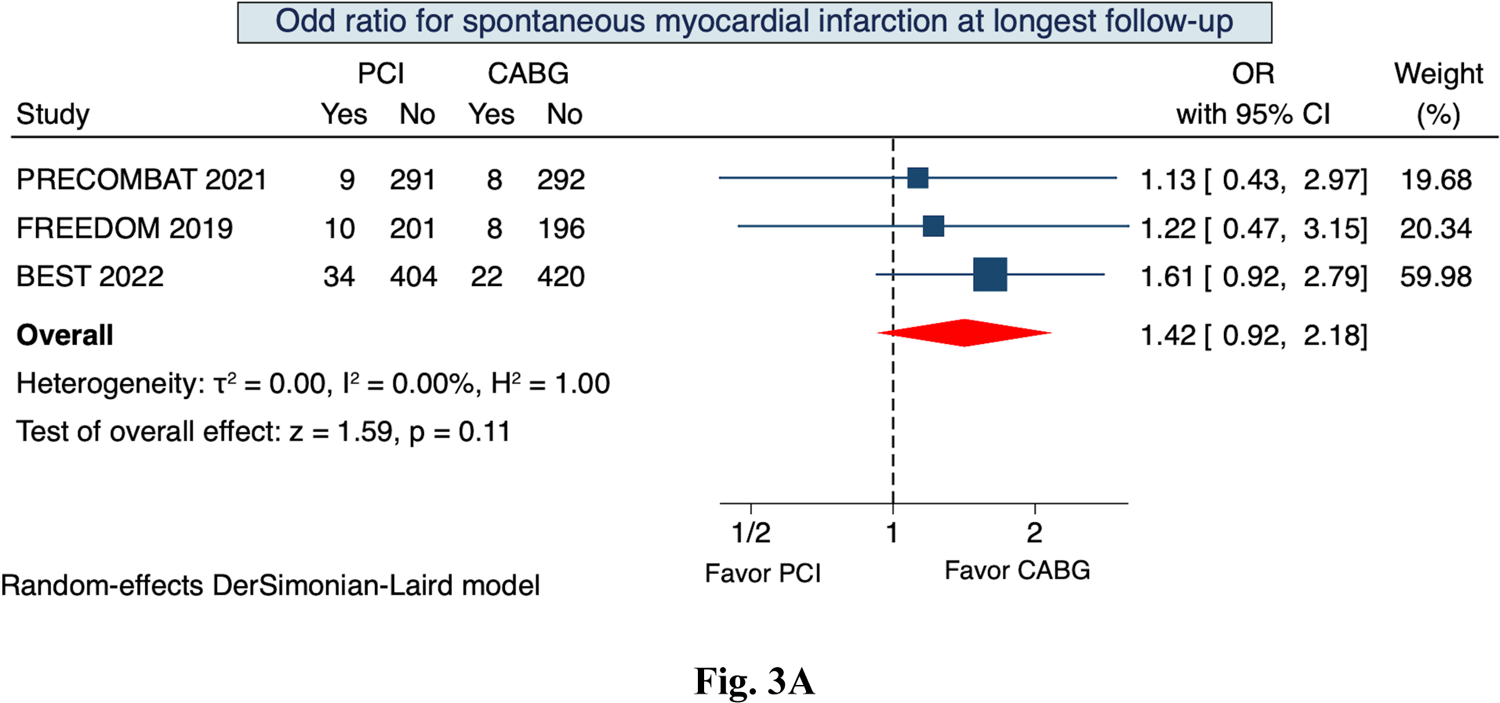
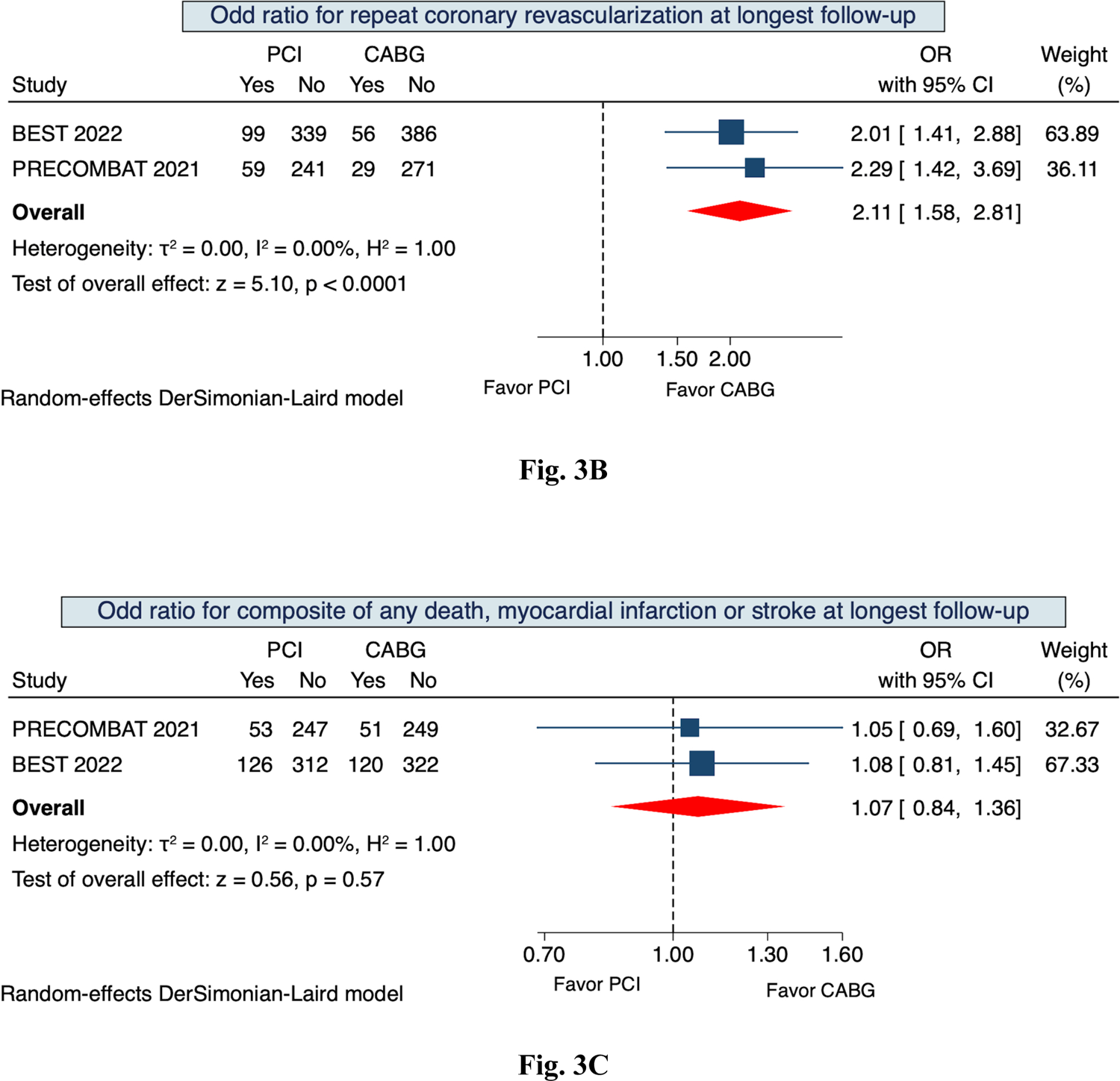
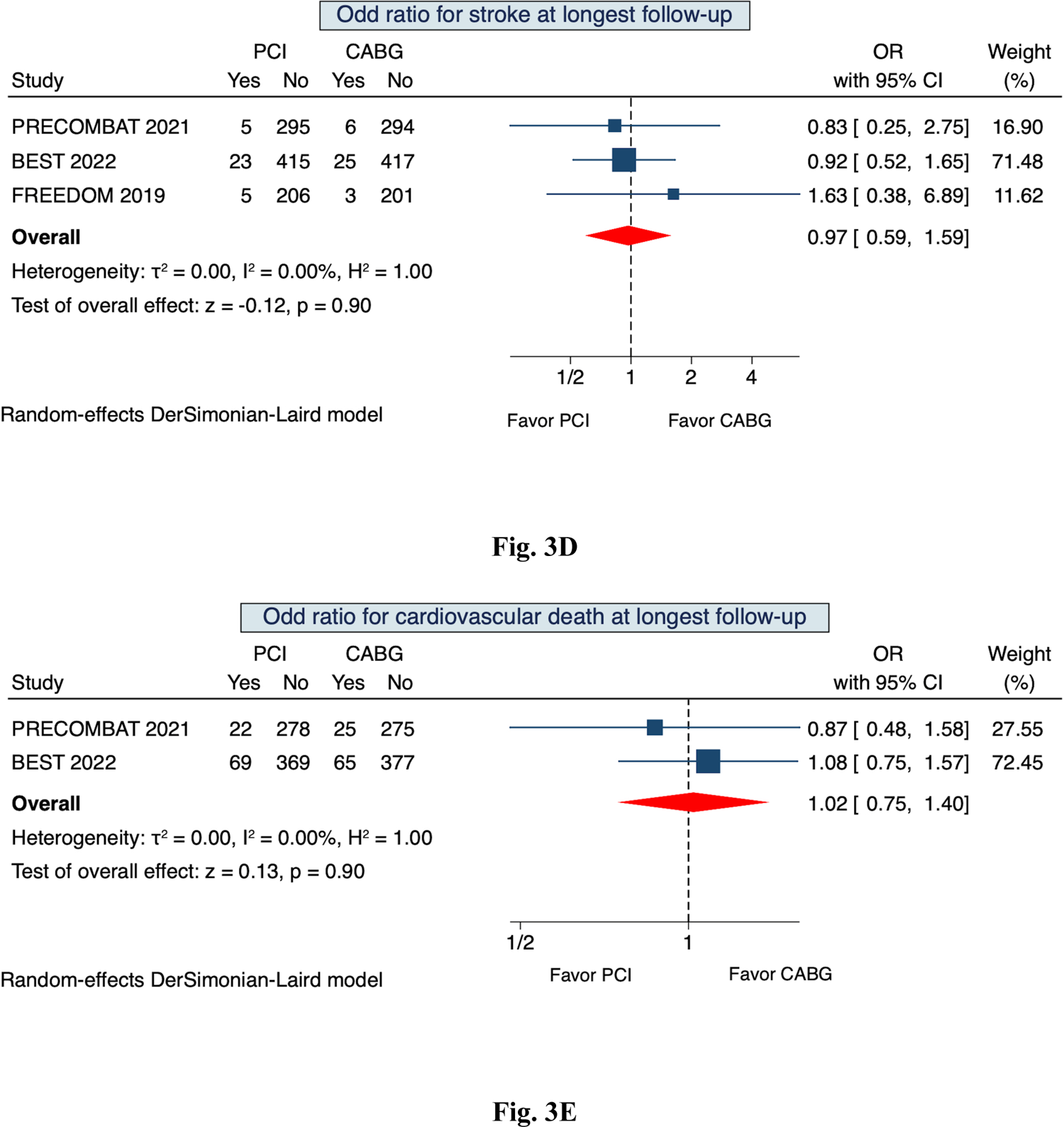
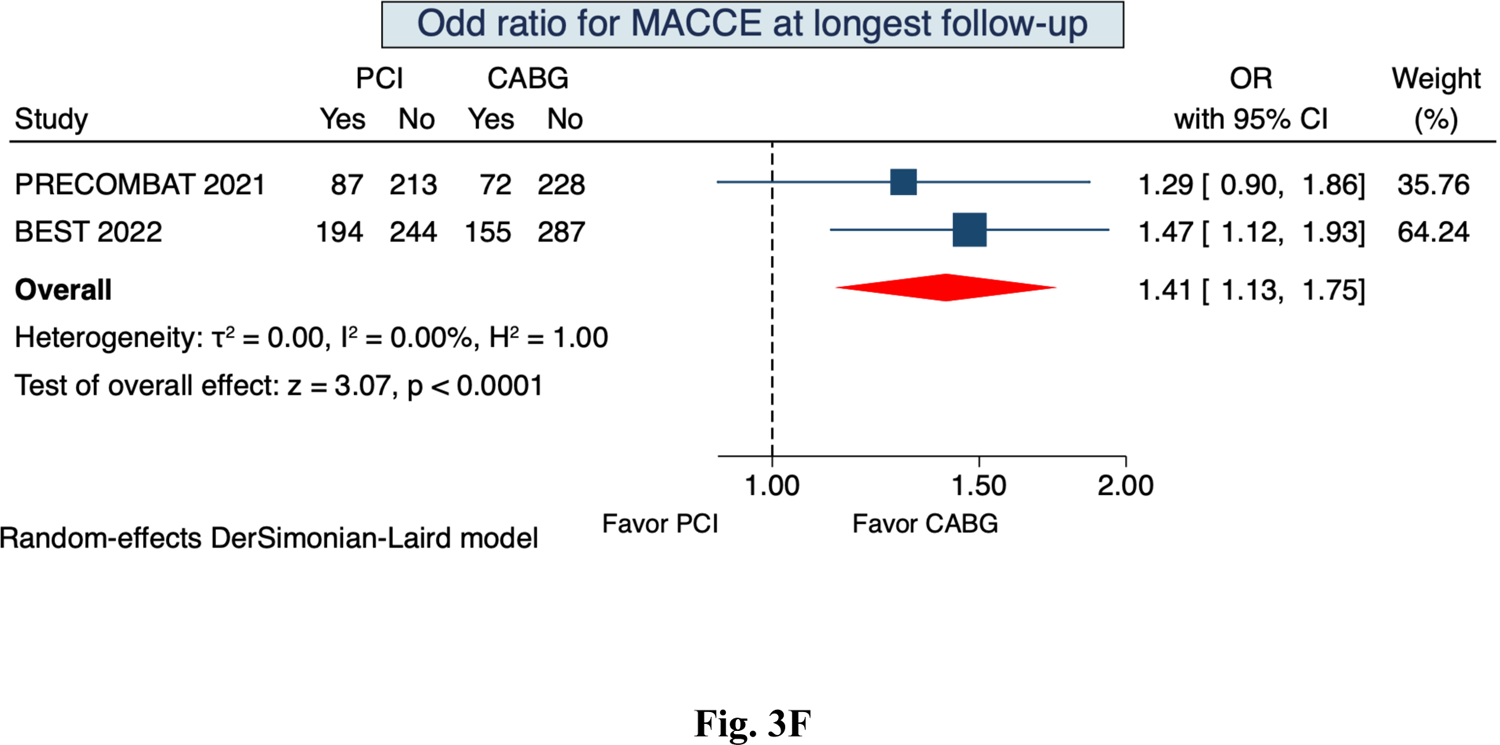
**(A)** Overall effect for myocardial infarction at longest follow-up following coronary artery bypass grafting (CABG) or percutaneous coronary intervention (PCI). **(B).** Overall effect for repeat coronary revascularization at longest follow-up following CABG or PCI. **(C)** Overall effect for composite of all-cause mortality, myocardial infarction or stroke at longest follow-up following CABG or PCI. **(D)** Overall effect for stoke at longest follow-up following CABG or PCI. **(E)** Overall effect for cardiovascular death at longest follow-up following CABG or PCI. **(F)** Overall effect for major adverse and cerebrovascular event (MACCE) at longest follow-up following CABG or PCI. *OR, odd ratio; CI, confidence interval;*

#### Repeat coronary revascularization

BEST and PRECOMBAT trials reported data for repeat coronary revascularization. The overall effect measure analysis showed that PCI was associated with a higher risk of repeat coronary revascularization (OR 2.11; 95% CI, 1.58-2.81; p<0.001) with no statistical heterogeneity (I^2^=0%) (Fig. 3B). Both trials reported the K-M curves. The time-to-event reconstructed curves with Cox linear regression are presented in supplementary material (Fig. S5).

#### Composite of all-cause mortality, stroke or MI

BEST and PRECOMBAT trials reported data for composite of all-cause mortality, stroke or MI. The overall effect measure analysis showed that PCI and CABG were comparable at longest follow-up (OR 1.07; 95% CI, 0.84-1.36; p=0.57) with no evidence of heterogeneity (I^2^=0%) (Fig. 3C). Both trials reported K-M curves for this composite outcome. The time-to-event reconstructed curves with Cox linear regression are presented in supplementary material (Fig. S6).

#### Stroke

Data on stroke beyond 5-year follow-up were reported in BEST, FREEDOM and PRECOMBAT trials. In the FREEDOM trial, data were recorded in 415 patients. The rate of stroke was comparable between the two interventions (OR 0.97; 95% CI, 0.59-1.59; p=0.91) without heterogeneity (I^2^=0%) (Fig. 3D).

#### Cardiovascular death

BEST and PRECOMBAT trials reported data for CV death. The overall effect measure analysis showed that PCI and CABG were comparable (OR 1.02; 95% CI, 0.75-1.40; p=0.90) with no heterogeneity (I^2^=0%) (Fig. 3E).

#### MACCE

BEST and PRECOMBAT trials reported data for MACCE. The PCI was associated with a higher rate of MACCE compared to CABG at available longest follow-up (OR 1.41; 95% CI, 1.13-1.75; p<0.0001) with no evidence of heterogeneity (I^2^=0%) (Fig. 3F).

## Discussion

The principal finding of this IPD study-level meta-analysis of RCTs is that, at a longest available 10-year follow-up, there was a significant greater risk of overall mortality after PCI. We estimated a 13.3% vs. 10.2% incidence of death after the first 5-years of PCI and CABG, respectively, and at 10-years the incidence of death was still higher in PCI (23.7% and 20.5%, respectively). However, this benefit was evident in the first 6-year follow-up; after this time, risk of death was similar between both interventions. We found that, at 10-year follow-up, patients undergoing surgery live longer and therefore CABG is the gold standard approach. However, this recommendation may not be as robust for younger patients or those with a longer life expectancy, because the benefit of CABG seems to be lost after 6 years of follow-up. At this point, survival curves stop diverging and become parallel. Therefore, for many patients, the decision between CABG or PCI should not be based on life expectancy, but on frailty or other considerations. Overall, we estimated a 2.4 months total gain in life expectancy in patients treated with CABG, compared to patients treated with PCI, indicating a favorable outcome of CABG. Interestingly, we also observed a progressively longer life expectancy in CABG compared with PCI, as revealed by analysis of RMST at 5, 8 and 10 years.

As previous IPD meta-analyses of RCTs (5,6,24) have already reported heterogeneous results of 5-year overall mortality in patients with LMD and/or MVD treated with CABG or PCI with DES, we thought it would be interesting to perform the landmark analysis at a cut-off time of 5 years, although the PHA not being violated. Interestingly, the incidence of all-cause mortality was significantly greater with PCI at the 0–5-year interval, while no difference was observed at the 5-10-year interval. The reasons for these different scenarios are multifactorial. One explanation could be related to the follow-up available in the FREEDOM trial, which reported data of 8-year follow-up including patients from only 25 of the 141 participating centers that agreed to participate in the extended follow-up study. Another explanation could be related to the CV-related death outcome that might be challenging to define even in RCTs, and therefore it could be plausible that CV death was not assigned appropriately in both interventions.

Several factors, including the extensive use of multiple arterial grafts (MAG) and performing graft anastomosis distal to the coronary stenosis, are associated with the significant reduction in mortality incidence in long-term follow-up. Several studies have reported a very high late survival rate (beyond 10-years) in patients who underwent CABG with MAG and these findings should be considered in Heart Team discussions (25–29). At the same time, advances in DES technology and increased adherence to dual antiplatelet therapy could really help to increasingly reduce the incidence of mortality and complications after PCI (30,31).

To the best of our knowledge, this study represents the first study-level IPD meta-analysis of RCTs focusing on follow-up beyond 5-years and has important clinical implications. Firstly, it includes trials with follow-up longer than 5-years and some previously unreported data. Secondly, we performed a reconstructed IPD meta-analyses curve to generate aggregated Kaplan–Meier plots and perform landmark analyses of primary endpoint. This allowed assessment of the entire follow-up duration of each RCT, and to calculate the overall RMST of primary endpoint for each intervention.

Based on the available data extracted to analyze the secondary endpoints at longest follow-up, the principal findings include a comparable incidence of MI, stroke, CV death and composite of death, MI or stroke. Interestingly, incidence of repeat coronary revascularization and MACCE were higher in PCI. Analysis of secondary endpoints is of great interest to guide Heart-Teams in making the most appropriate choice between one of the two revascularization interventions in those clinical scenarios where patients present with CAD potentially treatable with CABG or PCI. In such situations, the choice of one treatment over the other should also be guided by long-term data on the incidence of stroke, MI, repeat revascularization, and CV death. Unfortunately, the paucity of such data and the different definitions of the related outcomes adopted in the protocol of the included studies, did not permit to standardize the endpoints and have more complete and confident long-term results. Of note, the all-cause mortality primary outcome has an undeniable and identical definition in all trials, while the other outcomes may also suffer from a bias in event measure. For instance, data on secondary endpoints were reported extensively in BEST and PRECOMBAT trials, while FREEDOM trial reported a paucity data regarding the incidence of stroke and myocardial infarction. Therefore, we recognize that results of secondary endpoints of this meta-analysis should be considered with caution.

This meta-analysis has several strengths and limitations to be highlighted. As strengths, this is the first meta-analysis on this topic beyond 5-years follow-up. We reconstructed the databases and simulated a patient-level meta-analysis. This allowed us to graph the survival curves of the merged studies and accurately analyze the risks at certain cut-off points. Sample size beyond 5-years was more than 3700 patients. As limitations, firstly, all trials in the analysis had different inclusion and exclusion criteria; therefore, many patients were not included in the randomization according to the decision of the Heart Team of each trial. This might explain why patients with SYNTAX score > 33 were poorly represented in all trials. Secondly, individual trial reported different endpoint definition. While all-cause mortality is an unbiased measure outcome in all included trial, the other outcomes of interest were reported heterogeneously. Finally, the trials occurred in a period of more than 10 years and different DES technology and generations were used.

## Conclusion

In conclusion, in patients with LMD and or MVD there was a significantly overall higher incidence of all-cause mortality after PCI compared to CABG beyond 5-year follow-up. Specifically, CABG is still favorable beyond 5 years and maintains its gold standard role for the CAD treatment; PCI has an evident higher mortality during the first 5 years and a comparable outcome beyond 5 years. These results may help the Heart Team to tailor revascularization treatment and strategies for patients with LMD and MVD, favoring CABG for young patients and those with a life expectancy of at least 10 years, taking also into account that at 10 years after intervention, the life expectancy of CABG patients is significantly longer than that of PCI subjects by 2.4 months.

## Role of funding source

This meta-analysis was performed without funding.

## Data Availability

The data collection forms, data extracted from included studies and all other data and materials used in this meta-analysis will be available and sharable on specific and reasonable request to the corresponding author.

## Acknowledgement

The authors wish to thank Giorgia Pavan for her tireless contribution in English revision of the manuscript and Evelina Ceccato (medical librarian at University of Parma, Italy) for her precious support in literature search.

## Conflict of interest

none declared

## Abbreviations

CABG: coronary artery bypass grafting.

PCI: percutaneous coronary intervention

DES: drug-eluting stent

CAD: coronary artery disease

LMD: left main disease

MVD: multivessel disease

RCT: randomized controlled trial

OR: odd ratio

HR: hazard ratio

CI: confidence interval

RMST: restricted mean survival time

## Supplementary material

**Fig. S1.**
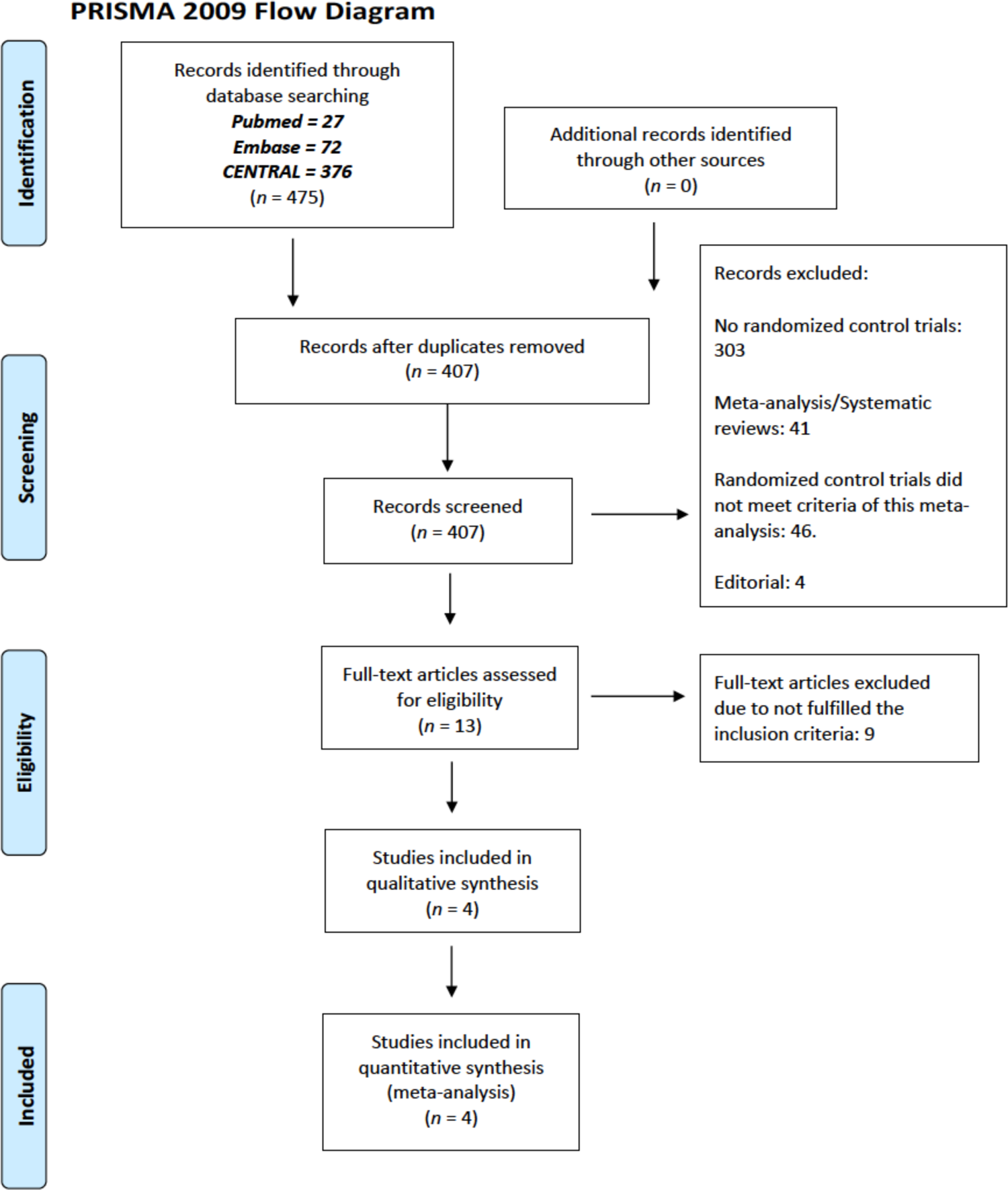
The PRISMA Flow Chart of study selection process

**Fig. S2.**
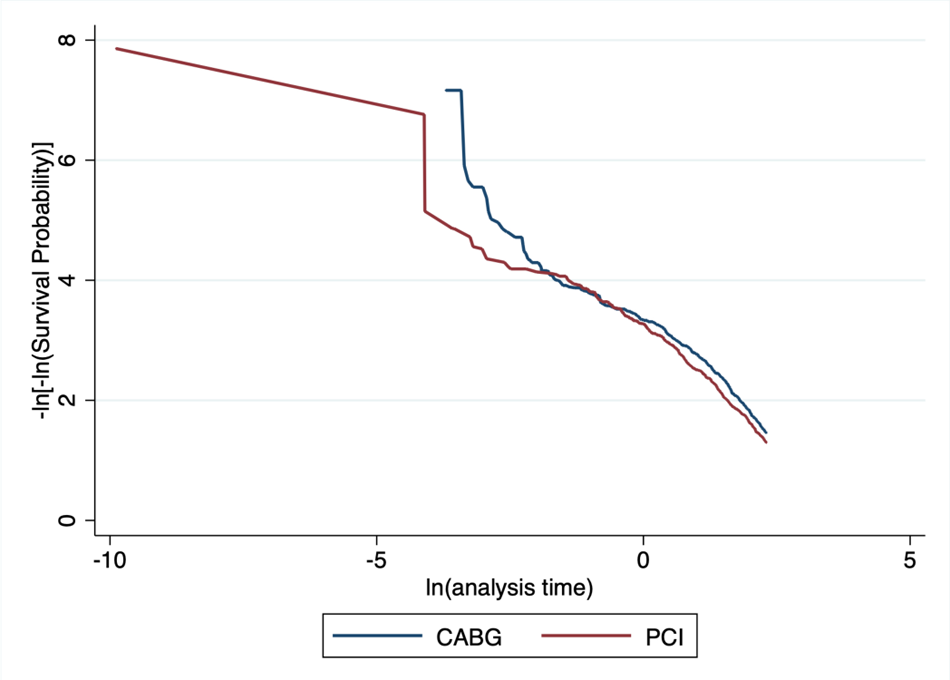
Log-minus-log survival curves

**Fig. S3.**
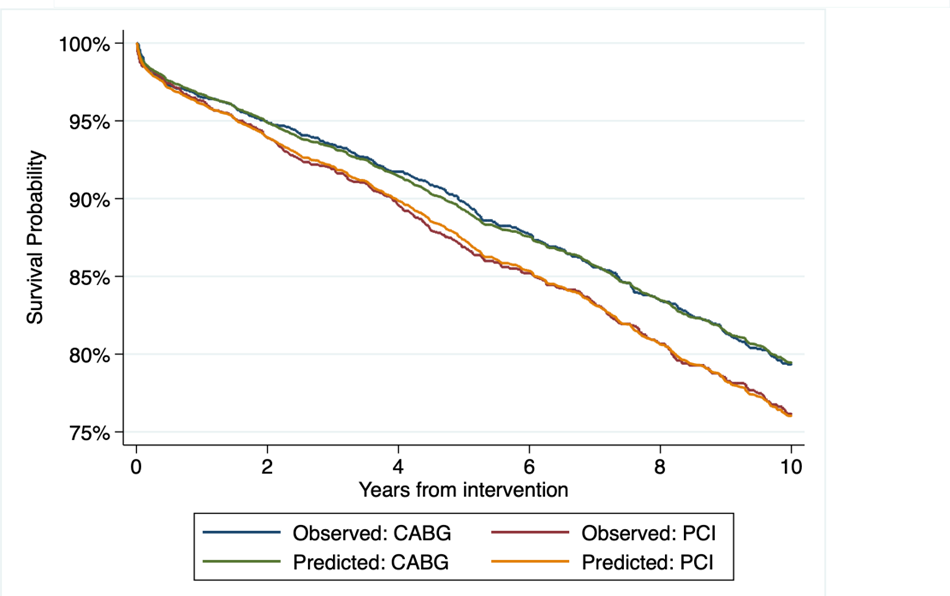
Predicted versus observed survival functions

**Fig. S4.**
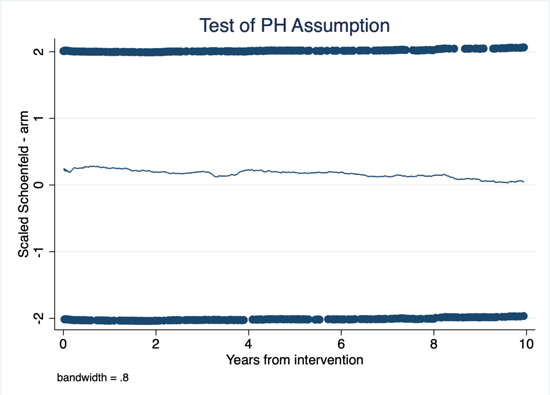
Scaled Schoenfeld residuals

**Fig. S5.**
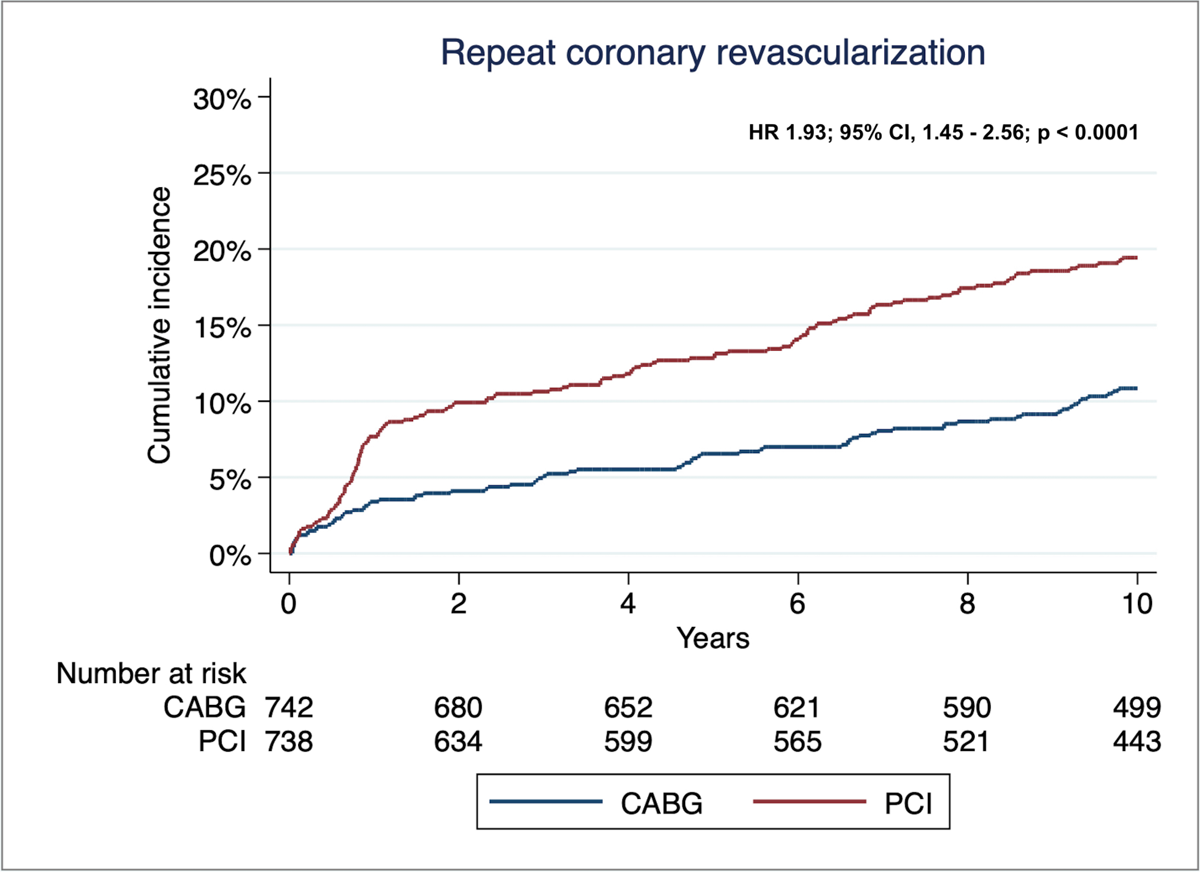
Time-to-event reconstructed curves for repeat coronary revascularization

**Fig. S6.**
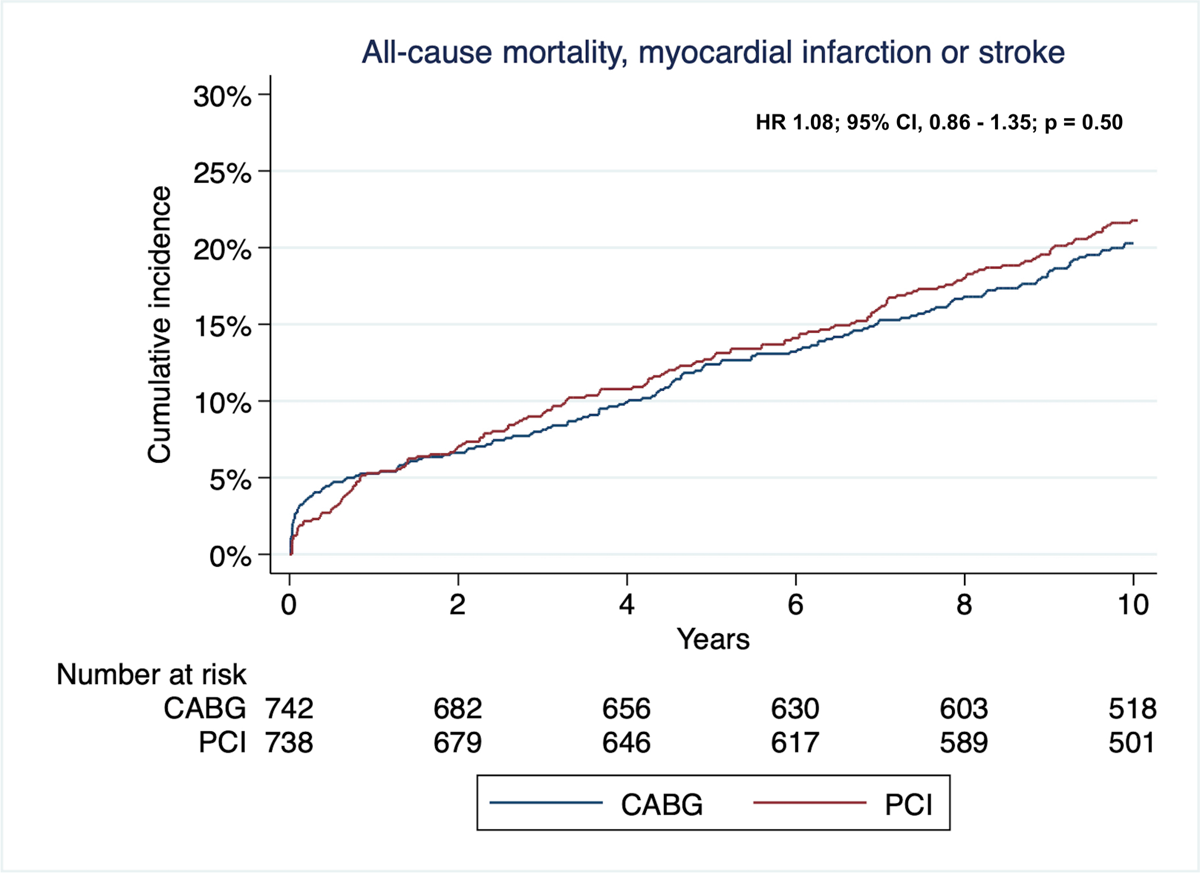
Time-to-event reconstructed curves for composite endpoints of all-cause mortality, myocardial infarction or stroke

**Table S1.** Algorithm literature search

| Search | PUBMED |  |
| --- | --- | --- |
| #1 | ((("coronary artery bypass"[Title/Abstract] OR "cabg"[Title/Abstract]) AND 2010/01/01:2023/01/31[Date - Publication]) AND (2010/1/1:2023/1/31[pdat])) | 23,119 |
| #2 | ((("drug eluting stent"[Title/Abstract] OR "des"[Title/Abstract] OR "stenting"[Title/Abstract]) AND 2010/01/01:2023/01/31[Date - Publication]) AND (2010/1/1:2023/1/31[pdat])) | 41,772 |
| #3 | ((("percutaneous coronary intervention"[Title/Abstract] OR "pci"[Title/Abstract]) AND 2010/01/01:2023/01/31[Date - Publication]) AND (2010/1/1:2023/1/31[pdat])) | 40,603 |
| #4 | ((("left main disease"[Title/Abstract] OR "left main coronary artery disease"[Title/Abstract]) AND 2010/01/01:2023/01/31[Date - Publication]) AND (2010/1/1:2023/1/31[pdat])) | 1,227 |
| #5 | ("multivessel coronary artery disease"[Title/Abstract] AND 2010/01/01:2023/01/31[Date - Publication]) AND (2010/1/1:2023/1/31[pdat]) | 914 |
| #6 | #1 AND #2 AND #3 AND #4 OR #5 | 1064 |
| #7 | ("randomised"[Title/Abstract] OR "randomized"[Title/Abstract] OR "trial"[Title/Abstract]) AND (2010/1/1:2023/1/31[pdat]) | 737,198 |
| #8 | ("long term follow up"[Title/Abstract] OR "extended follow up"[Title/Abstract]) AND (2010/1/1:2023/1/31[pdat]) | 39,605 |
| #9 | #7 AND #8 | 6,339 |
| #10 | #6 AND #9 | 27 |
| EMBASE |  |  |
| #1 | ('coronary artery bypass' OR 'cabg':ab,ti) AND [2010-2023]/py | 67,901 |
| #2 | ('drug-eluting stent' OR 'des' OR 'stenting':ti,ab) AND [2010-2023]/py | 373,598 |
| #3 | ('percutaneous coronary intervention' OR 'pci':ti,ab)<br>AND [2010-2023]/py | 108,182 |
| #4 | ('left main disease' OR 'left main coronary artery<br>disease':ti,ab) AND [2010-2023]/py | 2,511 |
| #5 | 'multivessel coronary artery disease':ti,ab AND [2010-2023]/py | 1,624 |
| #6 | (**randomized** OR **randomised** OR **trial**:ab,ti) AND [2010-2023]/py | 1,388,705 |
| #7 | #1 AND #2 AND #3 AND #4 | 449 |
| #8 | ('long term follow-up' OR 'extended follow-up' AND<br>[2010-2023]/py | 71,382 |
| #9 | #6 AND #7 AND #8 | **72** |
| **CENTRAL** |  |  |
| #1 | (left main disease):ti,ab,kw OR (left main coronary artery<br>disease):ti,ab,kw (Word variations have been searched) with<br>Publication Year from 2010 to 2023, in Trials | 1747 |
| #2 | (multivessel coronary artery disease):ti,ab,kw with<br>Publication Year from 2010 to 2023, in Trials (Word<br>variations have been searched) | 189 |
| #3 | (percutaneous coronary intervention):ti,ab,kw OR<br>("PCI"):ti,ab,kw (Word variations have been searched) with<br>Publication Year from 2010 to 2023, in Trials | 11849 |
| #4 | ("drug eluting stent"):ti,ab,kw OR (des):ti,ab,kw OR<br>(stenting):ti,ab,kw with Publication Year from 2010 to 2023,<br>in Trials (Word variations have been searched) | 13973 |
| #5 | ("coronary artery bypass"):ti,ab,kw OR (cabg):ti,ab,kw with<br>Publication Year from 2010 to 2023, in Trials (Word<br>variations have been searched) | 7608 |
| #6 | (randomized):ti,ab,kw OR (trial):ti,ab,kw OR<br>(randomised):ti,ab,kw with Publication Year from 2010 to<br>2023, in Trials (Word variations have been searched) | 902056 |
| #7 | #1 AND #3 AND #4 AND #5 | 226 |
| #8 | #7 OR #2 | 410 |
| #9 | #8 AND #6 | **376** |

**Table S2.** Risk of bias assessment using the Cochrane Collaboration revised tool for randomized control trials (RoB 2)

| Study | <u>D1</u> | <u>D2</u> | <u>D3</u> | <u>D4</u> | <u>D5</u> | <u>Overall</u> |
| --- | --- | --- | --- | --- | --- | --- |
| FREEDOM 2012 | Low risk | Low risk | Low risk | Low risk | Low risk | Low risk |
| SYNTAX 2013 | Low risk | Low risk | Low risk | Low risk | Low risk | Low risk |
| PRECOMBAT 2015 | Low risk | Low risk | Low risk | Low risk | Low risk | Low risk |
| BEST 2015 | Low risk | Low risk | Low risk | Low risk | Low risk | Low risk |
| EXCEL 2019 | Low risk | Low risk | Low risk | Low risk | Low risk | Low risk |
| NOBLE 2020 | Low risk | Low risk | Low risk | Low risk | Low risk | Low risk |

Low risk Some concerns High risk
- D1 Randomisation process - D2 Deviations from the intended interventions - D3 Missing outcome data - D4 Measurement of the outcome - D5 Selection of the reported result

**Table S3.**
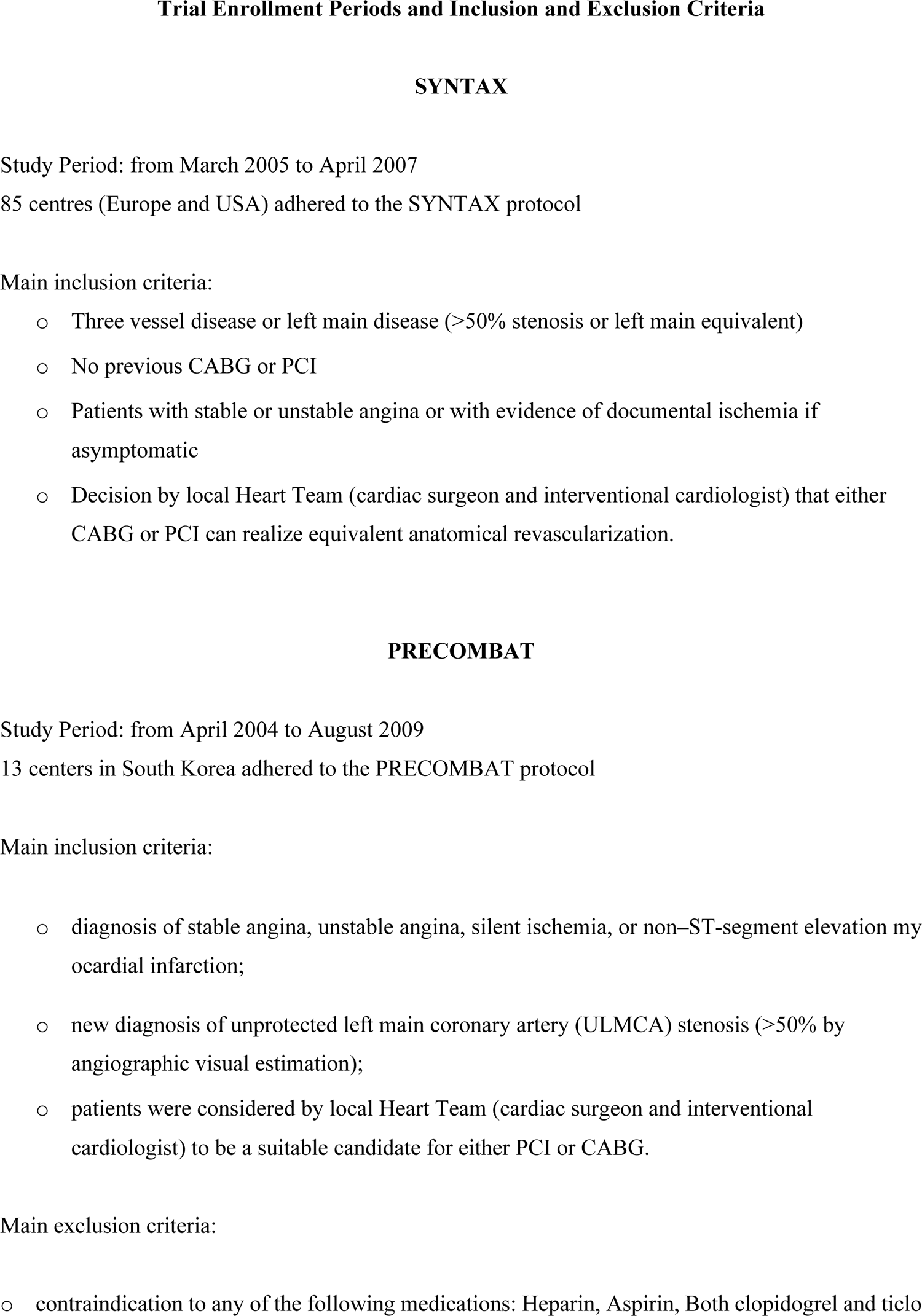
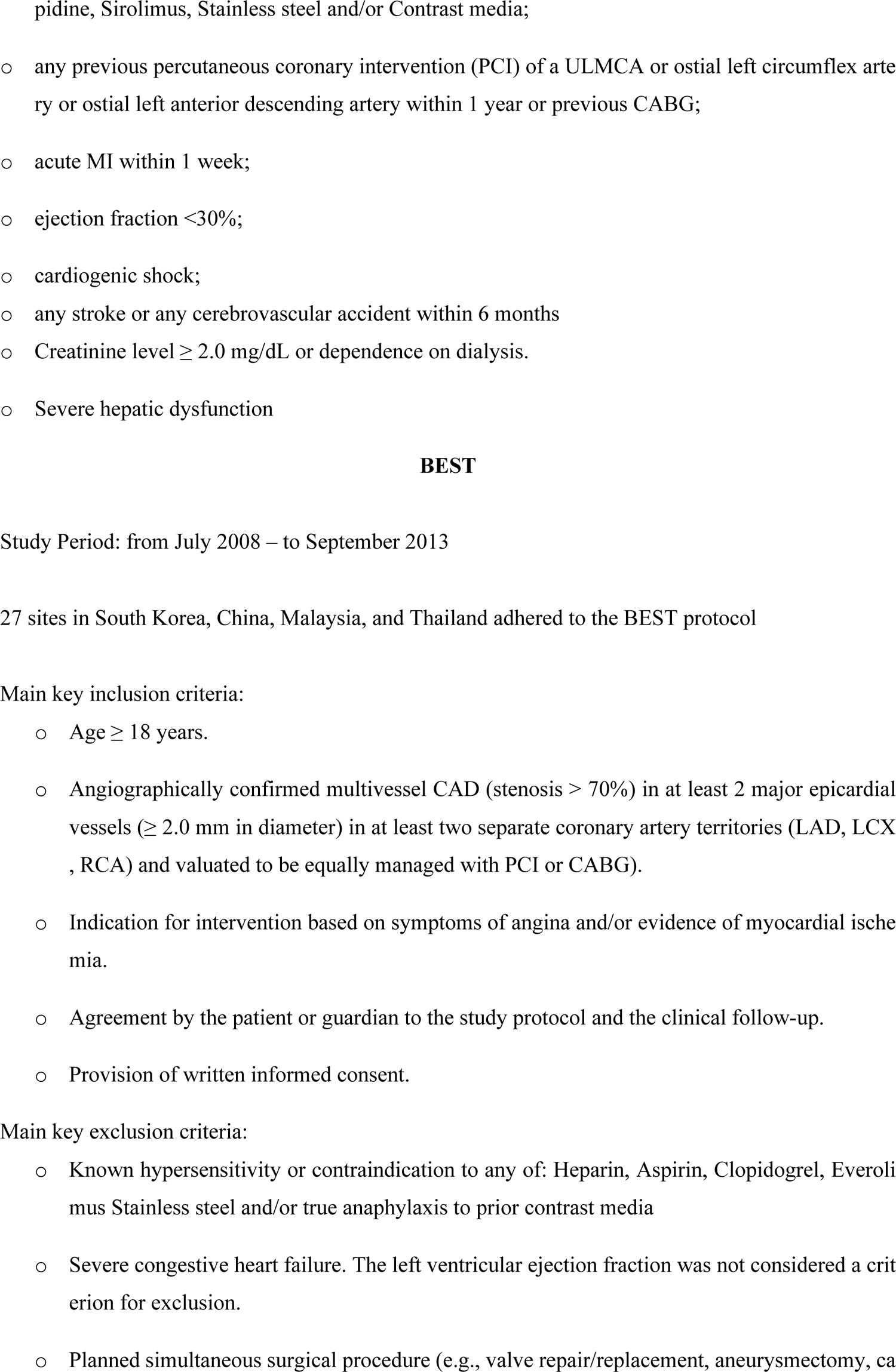
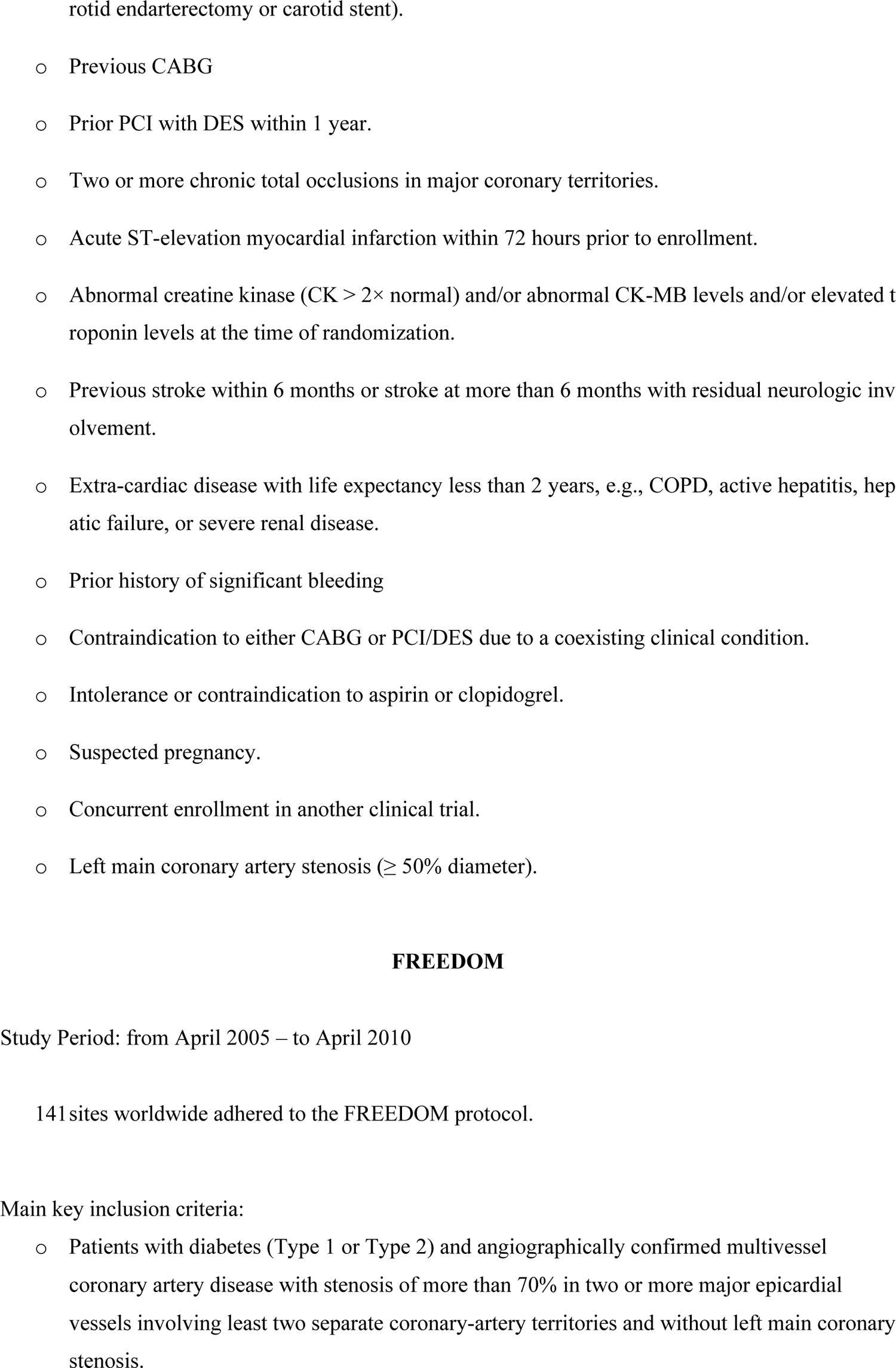
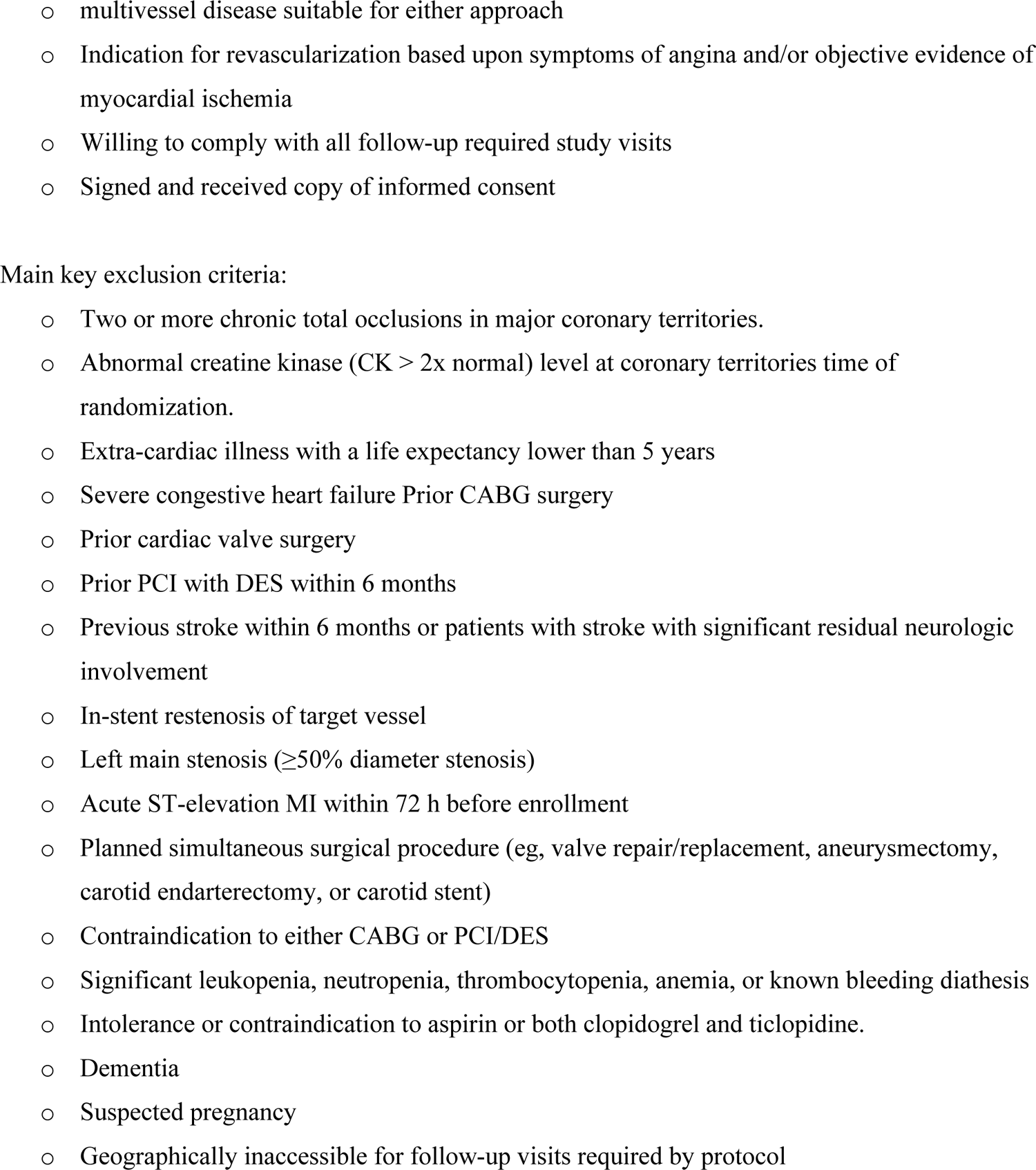
Endpoint definition of single trials

**Table S4.** Restricted Mean Survival Time (RMST)

| RMST | CABG | PCI | RMST difference | p-value |
| --- | --- | --- | --- | --- |
| 5-years | 4.71 (95% CI, 4.67-4.75) years | 4.64 (95% CI, 4.60-4.68) years | 0.07 years (25 days) (95% CI, 0.01-0.125;) | p=0.02 |
| 8-years | 7.16 (95% CI, 7.08-7.24) years | 7.30 (95% CI, 7.23-7.38) years | 0.14 years (1.7 months) (95% CI, 0.03-0.25;) | p=0.009 |
| 10-years | 8.93 (95% CI, 8.83-9.03) years | 8.73 (95% CI, 8.62-8.84) years | 0.20 years (95% CI, 0.05-0.35;) | p=0.007 |

## Notes

### Competing Interest Statement

The authors have declared no competing interest.

### Author Declarations

As the aggregated data were extracted from the published articles included in the analysis, this meta-analysis is exempted from Ethical Committee evaluation as the investigators of each trial obtained the approval from the local Ethical Committees.

